# Radiographic Assessment of Lung Edema (RALE) Scores are Highly Reproducible and Prognostic of Clinical Outcomes for Inpatients with COVID-19

**DOI:** 10.1101/2022.06.10.22276249

**Authors:** Nameer Al-Yousif, Saketram Komanduri, Hafiz Qurashi, Anatoliy Korzhuk, Halimat O. Lawal, Nicholas Abourizk, Caitlin Schaefer, Kevin J. Mitchell, Catherine M. Dietz, Ellen K. Hughes, Clara S. Brandt, Georgia M. Fitzgerald, Robin Joyce, Asmaa S. Chaudhry, Daniel Kotok, Jose D. Rivera, Andrew I. Kim, Shruti Shettigar, Allen Lavina, Christine E. Girard, Samantha R. Gillenwater, Anas Hadeh, William Bain, Faraaz A. Shah, Matthew Bittner, Michael Lu, Niall Prendergast, John Evankovich, Konstantin Golubykh, Navitha Ramesh, Jana J. Jacobs, Cathy Kessinger, Barbara Methé, Janet S. Lee, Alison Morris, Bryan J. McVerry, Georgios D. Kitsios

## Abstract

**INTRODUCTION:** Chest imaging is necessary for diagnosis of COVID-19 pneumonia, but current risk stratification tools do not consider radiographic severity. We quantified radiographic heterogeneity among inpatients with COVID-19 with the Radiographic Assessment of Lung Edema (RALE) score on Chest X-rays (CXRs).

**METHODS:** We performed independent RALE scoring by ≥2 reviewers on baseline CXRs from 425 inpatients with COVID-19 (discovery dataset), we recorded clinical variables and outcomes, and measured plasma host-response biomarkers and SARS-CoV-2 RNA load from subjects with available biospecimens.

**RESULTS:** We found excellent inter-rater agreement for RALE scores (intraclass correlation co-efficient=0.93). The required level of respiratory support at the time of baseline CXRs (supplemental oxygen or non-invasive ventilation [n=178]; invasive-mechanical ventilation [n=234], extracorporeal membrane oxygenation [n=13]) was significantly associated with RALE scores (median [interquartile range]: 20.0[14.1-26.7], 26.0[20.5-34.0] and 44.5[34.5-48.0], respectively, p<0.0001). Among invasively-ventilated patients, RALE scores were significantly associated with worse respiratory mechanics (plateau and driving pressure) and gas exchange metrics (PaO2/FiO2 and ventilatory ratio), as well as higher plasma levels of IL-6, sRAGE and TNFR1 levels (p<0.05). RALE scores were independently associated with 90-day survival in a multivariate Cox proportional hazards model (adjusted hazard ratio 1.04[1.02-1.07], p=0.002). We validated significant associations of RALE scores with baseline severity and mortality in an independent dataset of 415 COVID-19 inpatients.

**CONCLUSION:** Reproducible assessment of radiographic severity revealed significant associations with clinical and physiologic severity, host-response biomarkers and clinical outcome in COVID-19 pneumonia. Incorporation of radiographic severity assessments may provide prognostic and treatment allocation guidance in patients hospitalized with COVID-19.

## Introduction

Infection with the severe acute respiratory syndrome coronavirus-2 (SARS-CoV-2) has heterogeneous clinical presentations ranging from asymptomatic course to severe Coronavirus disease 2019 (COVID-19) with pneumonia and hypoxemia, requiring hospitalization. Inpatients with COVID-19 may require different levels of respiratory support, ranging from low level supplementation of inspired oxygen via nasal cannula in spontaneously breathing (SB) patients on the wards, to intubation and invasive mechanical ventilation (IMV) in the intensive care unit (ICU), to extracorporeal membrane oxygenation (ECMO) support in a selected subset of the sickest patients with refractory hypoxemia.

Multiple risk stratification tools for COVID-19 have been developed, combining clinical, physiologic, laboratory or research biomarker variables. Meanwhile, diagnosis of COVID-19 pneumonia relies on presence of radiographic consolidations on chest x-ray (CXR) or computed tomography (CT). Of the two modalities, CXR is the most widely available and routinely used, and CXRs are often repeated to determine pneumonia evolution or upon any new clinical indication^1,2^. However, radiographic severity has not been systematically integrated into risk predictions for COVID-19, and severity assessments are mostly qualitative and limited to narrative descriptions in diagnostic reports. The Radiographic Assessment of Lung Edema (RALE) score was developed and validated as a semi-quantitative instrument for evaluating the extent and density of radiographic opacities on CXRs in Acute Respiratory Distress Syndrome (ARDS).

RALE scores have been shown to correlate with severity of hypoxemia^3–5^, plasma biomarker levels (such as the soluble receptor of advanced glycation end-products - sRAGE)^6^, as well as to be prognostic of clinical outcome in non-COVID ARDS^3–5^. Nonetheless, individual studies analyzed small sets of ARDS subjects and CXRs, and associations with endpoints were inconsistent^6^. During the COVID-19 pandemic, RALE scores have been associated with COVID-19 pneumonia severity and clinical outcomes in several studies^7–10^, but we still lack a systematic evaluation of RALE scoring reproducibility and understanding of the impact of image-related variables (such as radiographic penetration) and patient covariates on derived RALE scores. Furthermore, it remains unknown whether RALE scores capture important inter-individual variability in clinical severity when examined in the context of provided respiratory support (e.g., intubated vs. non-intubated patients), and whether RALE scores reflect differences in underlying biological heterogeneity of COVID-19, as represented by host response biomarkers and subphenotypes, viral load or administered therapeutics.

In this study, we investigated the reproducibility of RALE scoring by multiple independent reviewers utilizing a dedicated software for image analysis and RALE score annotations. We analyzed CXRs in concert with detailed clinical and biological data from inpatients with COVID-19 enrolled in four independent cohort studies. We examined associations of RALE scores with cross-sectional indices of clinical severity, physiologic variables and biomarkers, and quantified the prognostic value of baseline RALE scores on COVID-19 clinical outcomes.

## Methods

### Discovery dataset

We analyzed data obtained from hospitalized patients with COVID-19, who were enrolled from April 2020 through October 2021 in one of three independent cohort studies within the UPMC Health System (detailed description available in the Supplement):

a. **The Acute Lung Injury Registry (ALIR) and Biospecimen Repository**, a prospective cohort study of critically ill adult patients (18-90 years of age) with acute respiratory failure. We enrolled COVID-19 subjects following admission to the ICU and obtaining informed consent (IRB protocol STUDY19050099), and collected plasma biospecimens.
b. **The COVID INpatient Cohort (COVID-INC)**, a prospective cohort study of moderately ill adult inpatients with COVID-19, hospitalized mainly in dedicated inpatient wards. Following informed consent (IRB protocol STUDY20040036), we collected blood biospecimens processed similarly to the ALIR study.
c. **The Prognostication for COVID-19 Patients Admitted to Intensive Care Units at UPMC Pinnacle (PROCOPI) study**, a retrospective cohort study of critically ill patients with COVID-19 hospitalized in ICUs at UPMC Pinnacle hospitals. We performed retrospective chart review and data collection (IRB protocol 20E059) for patients with COVID-19 on IMV.

### Clinical data collection

We extracted data on demographics, comorbid conditions, and clinical test results at baseline, and retrieved a portable CXR image. We scored each patient’s severity of illness according to the 10-point ordinal scale of the World Health Organization (WHO), and broadly classified baseline respiratory support in three categories: i) **SB** patients, i.e., not intubated subjects on various levels of oxygenation support including noninvasive ventilation, ii) **IMV**, intubated subjects in the ICU, and iii) **ECMO**, i.e., intubated subjects in the ICU on ECMO support. From IMV patients, we also collected detailed physiologic data from physician-set ventilatory parameters and obtained measurements for respiratory mechanics and gas exchange (Supplement), as previously described^11,12^. We recorded administered therapies and clinical endpoints across the COVID-19 timeline.

### RALE scoring

We performed RALE score assessments by ≥2 independent reviewers per image with the *Pulmo-Annotator* software (Veytel, LLC) (Figure 1 and details on scoring in the Supplement). In brief, we assessed radiographic penetration, image quality, presence of endotracheal tube, atelectasis, and then scored the most dense radiographic opacity in each quadrant by *extent* (scores of 0 for none, 1 for < 25%, 2 for 25–50%, 3 for 50–75% and 4 for > 75% of quadrant involved) and *density* (scores of 1 for hazy, 2 for moderate and 3 for dense consolidation). Each quadrant’s score was automatically calculated as the product of *extent*density*, and then all four quadrant scores were summed for a final RALE score (ranging from 0-48)^3^. Following a first iteration, each reviewer was provided feedback on scores distribution and agreement with other reviewer(s), followed by a joint session with the senior reviewer (GDK) to understand sources of disagreement and then independent re-scoring of CXRs with large discrepancies (a difference of score of ≥ 2 in any quadrant extent or density, or ≥15 RALE score difference). We used the RALE scores and annotated variables from the second iteration in quantitative analyses.

**Figure 1:**
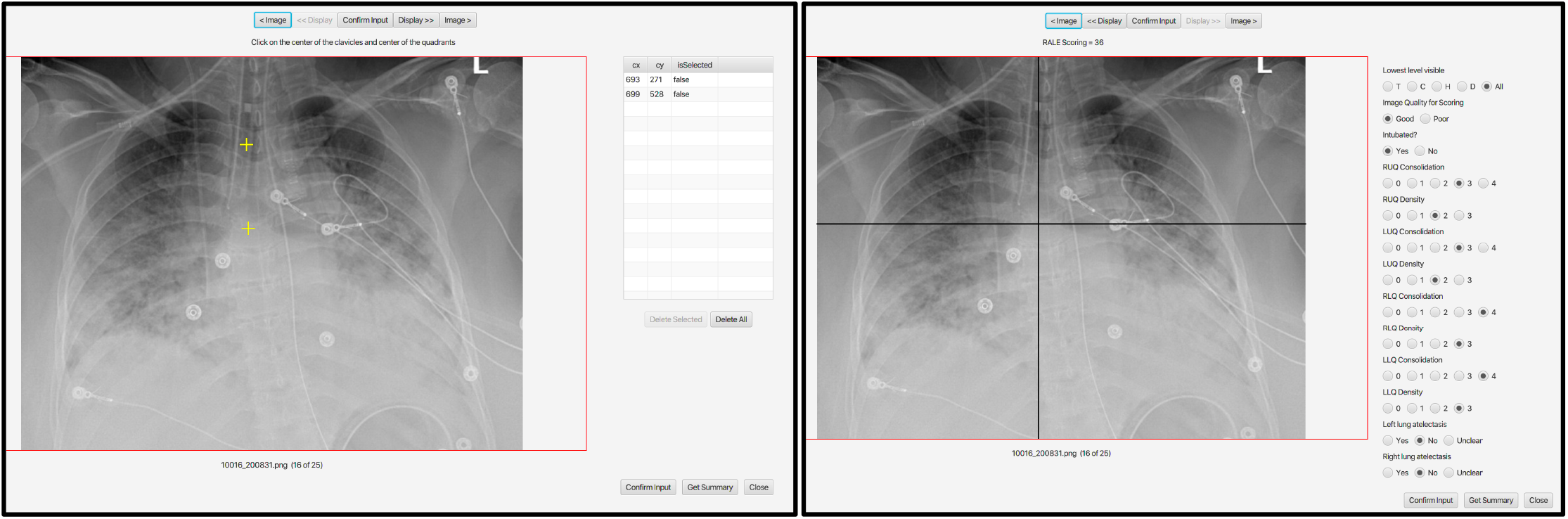
RALE scoring process through the *Pulmo-Annotator* software interface. This figure shows a screenshot of the *Pulmo-Annotator* software which was used to store and score the X-ray images. Left panel shows how the axis was set up with the first coordinate being assigned for the image rotation (vertical axis - mid-point of the vertebral column or mid-point of clavicles if image not rotated) and the second coordinate assigned for the horizontal axis determination at the level of the first branch of the left main bronchus. Right image shows the automated axes drawn by the software per the determined coordinates from the previous input, with options for physician annotation of image quality, penetration, presence of endotracheal tube, and atelectasis in each lung, and score options for density and consolidation extent for each quadrant, to allow for automated calculation of the final RALE score. Definition of abbreviations: RUQ = Right Upper Quadrant, LUQ = Left Upper Quadrant, RLQ = Right Lower Quadrant, and LLQ = Left Lower Quadrant.

### Plasma biomarkers

From available baseline samples from the ALIR and COVID-INC cohorts, we measured plasma biomarkers of injury and inflammation with custom-made Luminex panels as previously described^13^. We classified subjects into a hyperinflammatory vs. hypoinflammatory subphenotype by using predicted probabilities for subphenotype classifications from a published parsimonious logistic regression model utilizing interleukin-6 (IL-6), soluble tumor necrosis factor receptor 1 [sTNFR1] and bicarbonate^14^. In a random subset of plasma samples (n=63), we quantified circulating levels of SARS-CoV-2 RNA by qPCR, as previously described^15,16^.

### Statistical Analyses

We performed non-parametric comparisons for continuous (described as median and interquartile range – IQR) and categorical variables between clinical groups (Wilcoxon and Fisher’s exact tests, respectively). We examined for inter-reviewer agreement on RALE scores with Bland-Altman plots pre- and post-feedback sessions, and quantitatively by measuring inter-reviewer correlations and Intraclass Correlation Coefficients (ICC) in two-way random-effects models. For categorical variables on CXR assessments, we quantified inter-reviewer agreement with Cohen’s kappa statistics. We examined correlations of continuous variables with Pearson correlation test. We fit proportional hazards models to examine the statistical significance of baseline RALE scores on 60-day survival or time-to-liberation from IMV. We performed all analyses with the R software and a p-value of <0.05 was deemed statistically significant.

### Validation cohort

We obtained admission CXRs from 415 COVID-19 inpatients hospitalized within 18 different clinical sites of the Cleveland Clinic systems from March to October 2020. We collected clinical data from electronic medical records on demographics, comorbidities, physiologic and laboratory variables under an exempt review protocol (FLA 20-038) as previously described_17_. We classified patients into SB and IMV groups based on the type of respiratory support by the timing of the CXR.

All findings are reported in accordance with the STROBE statement for observational studies^18^.

## Results

### Characteristics of enrolled patients in the three discovery cohorts

We analyzed baseline CXRs from a total of 425 inpatients with COVID19 (154 subjects from ALIR, 138 from COVID-INC and 133 subjects from PROCOPI - [Table S1]) and stratified patients by level of respiratory support at time of the CXR as SB patients (n=178), IMV (n=234) and ECMO (n=13) (Table 1).

**Table 1.** Table 1: Baseline characteristics of subjects grouped by level of respiratory support: spontaneously-breathing patients (SB), patients on invasive mechanical ventilation (IMV) and patients on extracorporeal membrane oxygenation (ECMO).

|  | SB<br>(n = 178) | IMV<br>(n = 234) | ECMO<br>(n = 13) | p-value |
| --- | --- | --- | --- | --- |
| **Demographics** |  |  |  |  |
| Age, Years | 65.0 [55.0, 71.9] | 64.3 [55.1, 73.4] | 56.7 [50.7, 57.8] | 0.01 |
| Body Mass Index | 30.8 [26.1, 36.9] | 32.7 [28.5, 38.4] | 35.9 [32.4, 41.8] | 0.01 |
| Sex, Male | 84 (47.2%) | 160 (68.4%) | 10 (76.9%) | <0.01 |
| Race, White | 141 (79.2%) | 171 (73.1%) | 11 (84.6%) | 0.07 |
| Never Smokers | 77 (47.2%) | 90 (46.2%) | 7 (53.8%) | 0.86* |
| Resident of Nursing Facility | 21 (12.4%) | 29 (14.9%) | 0 (0.0%) | 0.28* |
| Diabetes Mellitus | 72 (40.4%) | 104 (44.4%) | 5 (38.5%) | 0.69 |
| Chronic Obstructive Lung Disease | 35 (19.7%) | 37 (15.8%) | 2 (15.4%) | 0.58 |
| Congestive Cardiac Failure | 24 (13.5%) | 35 (15.0%) | 0 (0.0%) | 0.31 |
| **Plasma biomarkers** |  |  |  |  |
| IL-6, pg/ml | 14.5 [5.9, 47.8] | 68.8 [14.0, 180.9] | 220.6 [88.1, 1112.0] | <0.01 |
| IL-8, pg/ml | 16.0 [8.6, 29.7] | 21.7 [14.2, 42.8] | 27.8 [15.0, 46.8] | <0.01 |
| Ang2, pg/ml | 2701.7 [1426.7, 4090.9] | 5634.2 [2860.3, 10913.3] | 5927.3 [4170.2, 7592.2] | <0.01 |
| Procalcitonin, pg/ml | 107.5 [65.0, 283.9] | 633.7 [173.3, 1956.3] | 465.6 [196.6, 1261.5] | <0.01 |
| ST2, ng/ml | 87.8 [47.3, 16.8] | 211.1 [96.3, 378.9] | 190.1 [111.0, 253.9] | <0.01 |
| Pentraxin-3, pg/ml | 5791.2 [2621.5, 11728.4] | 9599.2 [4680.4, 21226.8] | 5525.4 [3748.7, 11688.9] | 0.01 |
| sRAGE, pg/ml | 3979.1 [2447.0, 9137.2] | 6158.5 [3269.3, 12653.0] | 1754.8 [1219.6, 5102.5] | <0.01 |
| sTNFR1, pg/ml | 3438.2 [2485.5, 5359.9] | 5601.7 [3375.9, 12297.4] | 7283.8 [4650.7, 8515.4] | <0.01 |
| Hyperinflammatory Phenotype | 5 (3.5%) | 12 (16.2%) | 2 (16.7%) | <0.01* |
| **Radiographic parameters** |  |  |  |  |
| RALE Score, total | 20.0 [14.1, 26.7] | 26.0 [20.5, 34.0] | 44.5 [34.5, 48.0] | <0.01 |
| Lower Quadrants RALE Score | 14.0 [10.0, 17.0] | 16.0 [12.5, 20.0] | 24.0 [22.0, 24.0] | <0.01 |
| Upper Quadrants RALE Score | 6.0 [3.5, 9.9] | 10.0 [7.0, 14.0] | 22.5 [12.8, 24.0] | <0.01 |
| Time of CXR from symptom onset | 7.0 [3.0, 11.0] | 9.0 [6.0, 15.0] | 14.0 [11.0, 22.0] | <0.01 |
| **Clinical outcomes** |  |  |  |  |
| In-Hospital Mortality | 33 (18.5%) | 163 (69.7%) | 6 (46.2%) | <0.01 |
| **Discharge destination** |  |  |  |  |
| Home | 113 (63.5%) | 17 (7.3%) | 1 (7.7%) | <0.01 |
| Inpatient Rehabilitation | 2 (1.1%) | 15 (6.4%) | 2 (15.4%) | <0.01 |
| Long Term Acute Care | 3 (1.7%) | 18 (7.7%) | 3 (23.1%) | <0.01 |
| Skilled Nursing Facility | 27 (15.2%) | 21 (9.0%) | 1 (7.7%) | 0.154 |

Continuous variables are reported in median [interquartile range]. Categorical variables are reported as n (%).
\* = we only included patients with available clinical data or research biomarkers in analysis. Patients with unavailable data were excluded.
Definition of abbreviations: CXR = Chest X-Ray, IL = Interleukin, Ang2 = Angiopoietin-2, ST2 = Suppression of Tumorigenicity-2, sRAGE = Soluble Receptor of Advanced Glycation End-Products, NMBA = Neuromuscular Blocking Agents, ECMO = Extracorporeal Membrane Oxygenation, IPR = Inpatient Rehab, LTAC = Long Term Acute Care, and SNF = Skilled Nursing Facility.

### Inter-rater agreement for RALE scores

In first iteration of RALE scoring, we found good inter-rater agreement between reviewers for total RALE scores (ICC 0.85, 95% confidence interval-CI [0.82-0.88], p<0.0001), with 18/425 (4%) of CXRs showing large RALE score discrepancies (+/-15 points) between two reviewers. Following feedback, the inter-rater agreement on RALE scores at the second scoring iteration improved to excellent (ICC 0.93 [0.92-0.95], p<0.0001), with 4/425 (<1%) CXRs showing large RALE discrepancies (Figure 2 and Tables S2-3). We then used average RALE scores from two reviewers in further quantitative analyses.

**Figure 2:**
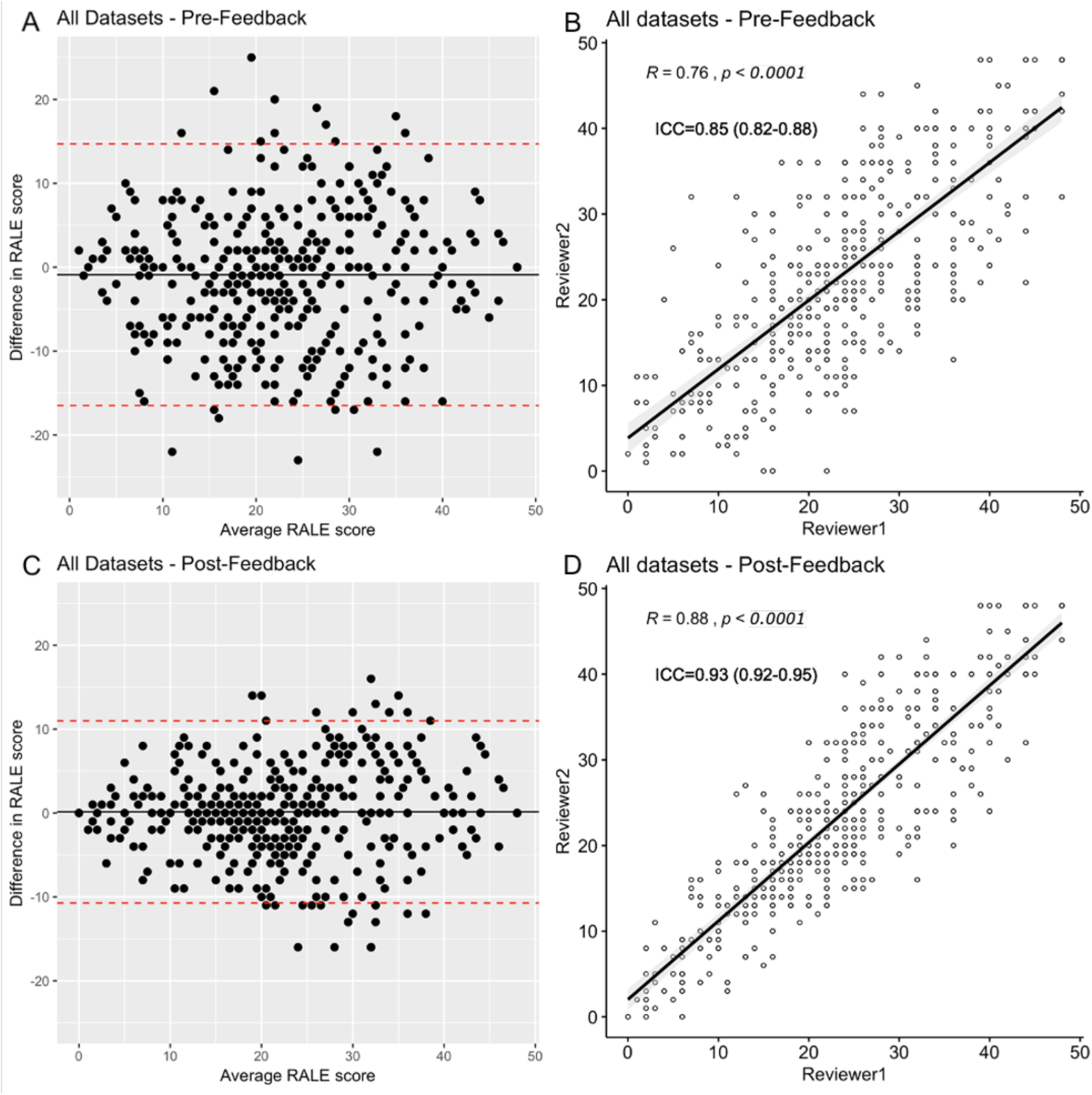
Inter-rater agreement for RALE scores in pre- and post-feedback session assessments. Top figures show pre-feedback results of the inter-rater agreement. The left upper panel image (A) shows a Bland–Altman plot and the right image (B) shows a scattered plot graph with a high correlation coefficient (R = 0.76, p <0.0001) and good inter-rater agreement (intraclass correlation coefficient - ICC 0.85 [0.82-0.88], p <0.0001) between the reviewers. Pre-feedback, 4% of CXRs had large RALE score discrepancies (+/- 15 points). The bottom panels show post-feedback results of the inter-rater agreement. The left image (C) shows a Bland–Altman plot and the right image (D) shows a scattered plot graph with a very high correlation coefficient (R = 0.88, p <0.0001) and excellent inter-rater agreement (ICC 0.93 [0.92-0.95], p <0.0001) between the reviewers. Post-feedback, less than 1% of images had large RALE score discrepancies.

### Impact of CXR image variables on RALE scores

We examined for the association between CXR image findings and RALE scores without any knowledge of clinical data. Under-penetrated CXRs (i.e., CXRs in which vertebral bodies were visible only behind the trachea) had higher median RALE scores compared to CXRs with visible vertebral bodies behind the heart (p<0.01, Figure S1), and right lung atelectasis (definite or possible) was associated with significantly higher scores for right lower quadrant mean density scores (p<0.01, Fig S1). Overall, the lower quadrants (right and left) had much higher quadrant scores compared to their corresponding upper quadrants (right and left, respectively, p<0.0001). Left lower quadrant scores were statistically significantly higher than right lower quadrant ones (p<0.01, Fig S1). Therefore, both radiographic penetration and physician-ascribed presence of atelectasis were shown to have an impact on RALE scores, with the lower quadrant scores being systematically higher than the upper quadrants.

### RALE scores by baseline level of respiratory support and period of the pandemic

ECMO patients had the highest RALE scores (median [IQR]: 44.5 [34.5-48.0]), followed by IMV (26.0 [20.5-34.0]) and then by SB patients (20.0 [14.1-26.7]), p<0.0001) (Figure 3A). The association between radiographic and clinical severity was also significant for the component RALE scores in each quadrant (Figure 3B-C) and by WHO ordinal scale categories (Figure 3D). The COVID-INC cohort had the highest proportion of SB patients (91%) and as expected, patients in the COVID-INC cohort had lower RALE scores compared to the ALIR and PROCOPI cohorts (p<0.0001, Figure S2A). Throughout the period of enrollment (March 2020-October 2021), we found that there was a progressive increase of baseline RALE scores over the epoch of time for IMV patients only (R=0.16 for RALE scores and time from March 2020 till CXR date, p=0.017, Figure S2B).

**Figure 3:**
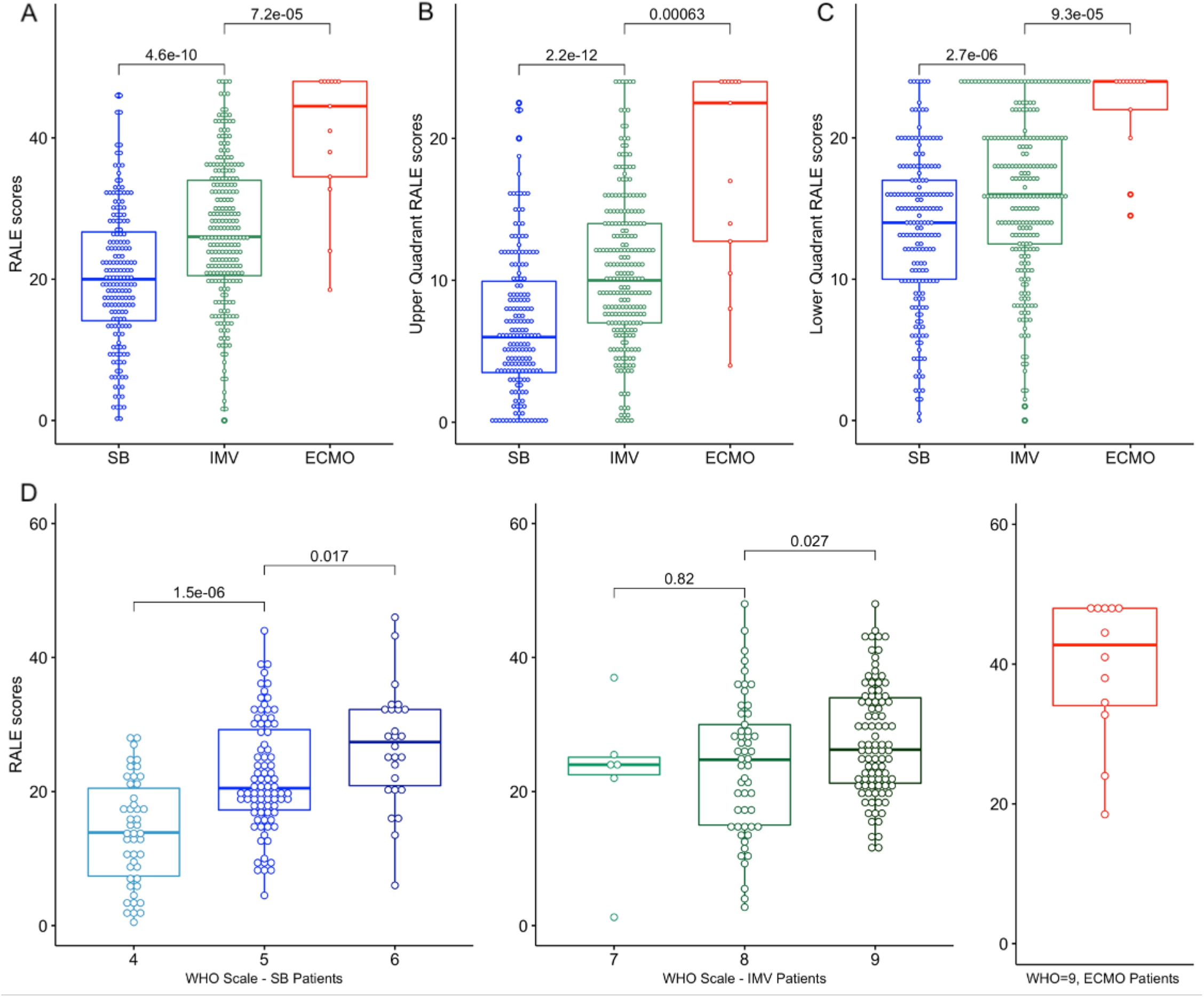
Patients on higher levels of respiratory support had higher RALE scores at baseline. A: Patients on ECMO had much higher RALE scores compared to patients on IMV, who in turn had significantly higher RALE scores than SB patients. B and C: We found similar differences in upper and lower quadrant RALE scores by level of respiratory support. D. Total RALE scores were significantly higher by rising disease severity based on the ordinal WHO scale. Definition of abbreviations: WHO = World Health Organization, and ECMO = Extracorporeal Membrane Oxygenation

### Baseline clinical variables and RALE scores

We then examined for associations between clinical characteristics and RALE scores at baseline, separately for SB, IMV and ECMO patients, given the significantly different RALE scores by respiratory support category. Among SB patients, men and obese patients had higher RALE scores (p<0.05, Figure S3) whereas among IMV patients, nursing facility residents and patients with history of Chronic Obstructive Pulmonary Disease (COPD) had significantly lower RALE scores than their counterparts (p<0.0001, Figure S3). Notably, for patients on IMV, age was inversely correlated with RALE scores (p<0.0001), whereas for both SB and IMV patients RALE scores were positively correlated with body mass index (BMI, p<0.0001) and duration of COVID-19 symptoms (p<0.0001) (Figure S4).

### Pulmonary physiology and applied therapies are associated with RALE scores

We examined physician-set ventilatory parameters, pulmonary mechanics and gas exchange metrics in IMV patients only, because such measurements are either unavailable or not reliably measured in SB patients, and confounded by the extracorporeal support in ECMO patients. In terms of ventilatory parameters, RALE scores were inversely correlated with set tidal volumes (TV, R=-0.17, p=0.02), and were higher by increasing levels of positive end-expiratory pressure (PEEP, Figure 4A-B). By measured mechanics, RALE scores positively correlated both with plateau (R=0.38, p<0.0001) and driving pressures (R=0.31, p<0.001, Figure 4C-D). For gas exchange, RALE scores were positively correlated with ventilatory ratios (i.e., worse CO2 clearance, R=0.18, p=0.02) and negatively correlated with PaO_2_/FiO_2_ ratios (i.e., worse hypoxemia, R=-0.3, p<0.0001, Figure 4E-F). Patients on IMV who underwent prone positioning or received neuromuscular blockade had higher RALE scores than their untreated counterparts (p<0.0001, Figure S5).

**Figure 4:**
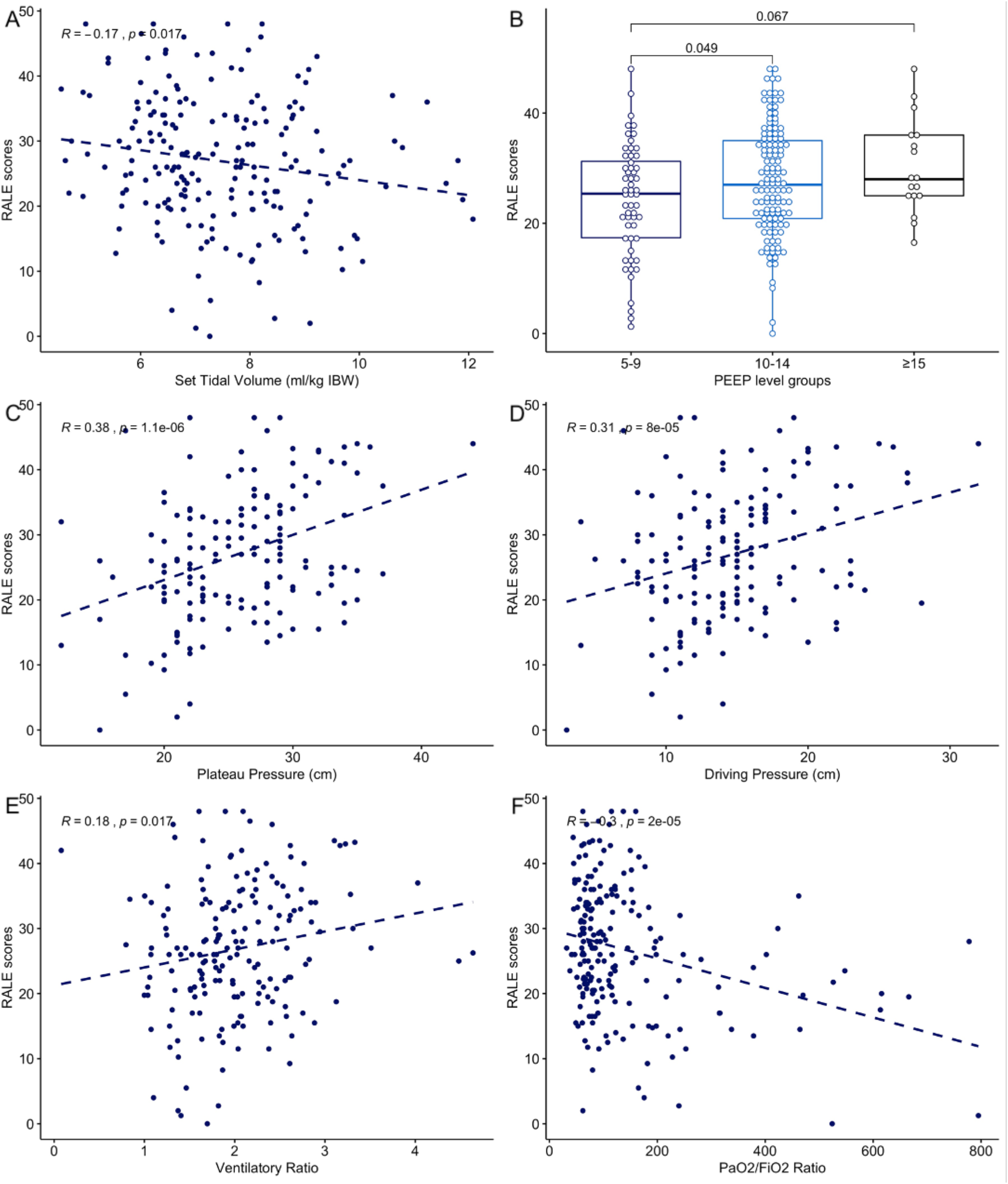
RALE scores were significantly associated with pulmonary dysfunction metrics in invasively mechanically ventilated patients. RALE scores were significantly associated with physician set parameters on mechanical ventilation (A: tidal volumes expressed in ml/kg of ideal body weight; B: RALE scores by positive end-expiratory pressure levels), correlated with respiratory mechanics (C: plateau pressure and, D: driving pressure) and gas exchange parameters (E: positive correlation with ventilatory ratio, i.e. worse CO2 clearance and F: inverse correlation with PaO2/FiO2 ratio, i.e. worse hypoxemia). Definition of abbreviations: PEEP = Positive End Expiratory Pressure, PaO2 = partial pressure of oxygen, and FiO2 = Fraction of Inspired Oxygen.

### RALE scores and plasma biomarkers

We did not examine plasma biomarker associations in ECMO patients due to small sample size. We found no significant association between RALE scores and plasma SARS-CoV-2 RNA levels (“viral RNA-emia”) in either SB or IMV patients examined separately. Baseline RALE scores correlated significantly with plasma levels of IL-6 in SB patients, and with IL-6, sTNFR1 and sRAGE levels in IMV patients (Figure 5A-B). When stratified into subphenotypes, hyper-inflammatory patients had higher RALE scores in both SB patients (p=0.04) and IMV patients (p=0.007, Fig 5C-D).

**Figure 5:**
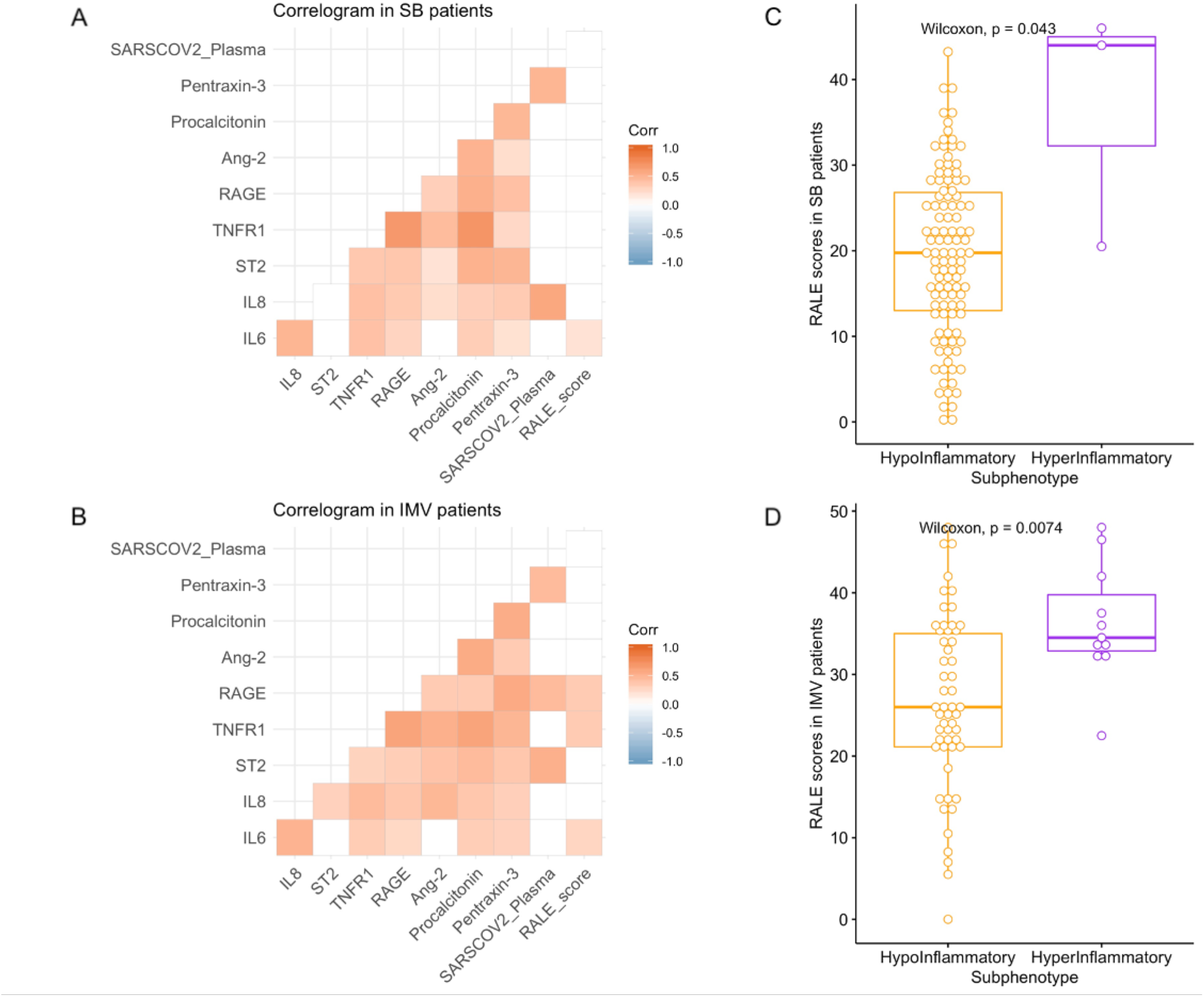
RALE scores were significantly correlated with biomarkers of host injury and inflammation, and significantly associated with the hyperinflammatory subphenotype. A-B: correlograms of host response biomarkers, SARS-CoV-2 viral load and RALE scores in SB (A) and IMV (B) patients. Pearson’s correlations are shown in color code (red for positive and blue for negative correlations) and only statistically significant correlations following adjustment for multiple testing by Benjamini-Hochberg method are shown. C-D: Patients assigned to the hyperinflammatory subphenotype (based on a prediction from a parsimonious predictive model utilizing IL-6, sTNFR1 and bicarbonate levels) had higher RALE scores in both SB (C) and IMV (D) patients. Definition of abbreviations: IMV = Invasive Mechanical Ventilation, IL = Interleukin, ST2 = Suppression of Tumorigenicity-2, sTNFR1 = Soluble Tumor Necrosis Factor Receptor 1, sRAGE = Soluble Receptor of Advanced Glycation End-Products, and Ang2 = Angiopoietin-2.

### RALE scores are prognostic of clinical outcomes

When all patients were combined (SB, IMV, ECMO), baseline RALE scores were higher among non-survivors (25.1 [19.8-33.0]) compared to survivors of hospitalization (22.3 [15.0-31.0], p=0.0014, Figure 6A). In a Cox proportional hazards model for 60-day survival adjusted for age, sex, BMI and COPD, RALE scores were significantly associated with worse survival (adjusted hazard ratio 1.02 [1.01-1.04] for each unit increase in RALE score, p=0.002). Stratified by RALE score tertiles (low <19.6, intermediate: 19.6–28.5, high >28.5), patients in the high tertile had worse 60-day survival by Kaplan-Meier curve analysis (Figure 6B). When examined separately within each group of respiratory support level, RALE scores were not significantly associated with 60-day survival in adjusted Cox proportional hazards models. Similarly, we did not find a significant association for RALE scores with time to liberation from IMV in Cox models adjusted for age, sex, BMI, COPD, TV and PEEP levels.

**Figure 6:**
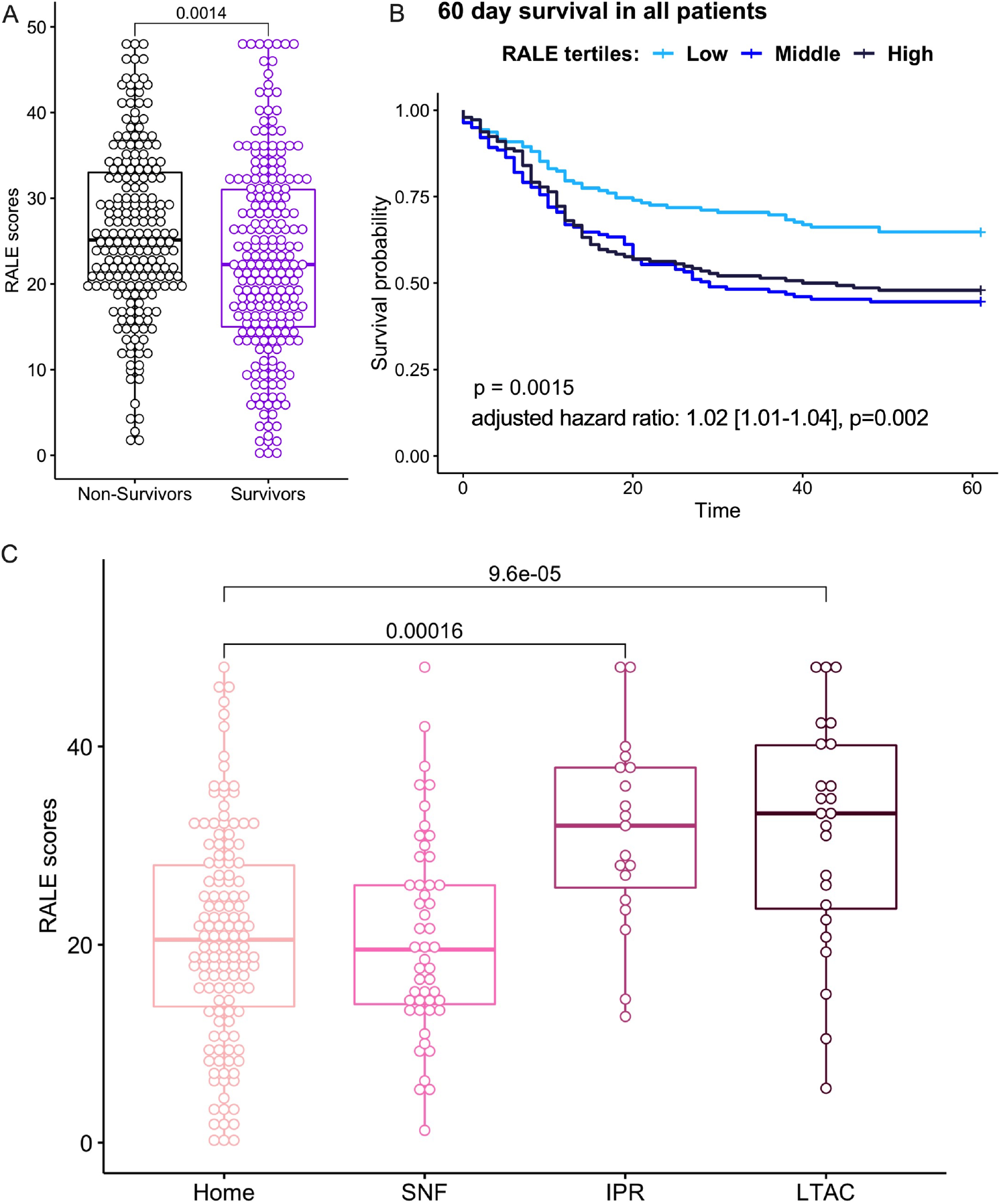
RALE score association with clinical outcomes. A: Non-survivors had higher RALE scores than survivors (25.1 [19.8-33.0] vs 22.3 [15.0-31.0], p = 0.0014). B. By Kaplan-Meier curve analysis, patients in the low tertile of RALE scores (<19.6) had improved survival compared to patients in middle/high tertiles. C. Higher care needs on final disposition were associated with higher RALE scores at baseline: long-term acute care facilities (LTAC) (33.3 [22.9-40.4]) or In-Patient Rehab (IPR) (32.0 [24.5-38.0]), vs. skilled nursing facility (SNF) (19.5 [13.9-27.3]) or home care (20.5 [13.5-28.0], p <0.0001).

Among survivors of hospitalization, higher complexity of care needs on discharge (based on disposition destination) were significantly associated with baseline RALE scores, with higher RALE scores for survivors discharged to a long-term acute care facility (33.3 [22.9-40.4]) or in-patient rehabilitation (32.0 [24.5-38.0]) compared to those discharged to a skilled-nursing facility (19.5 [13.9-27.3]) or home care (20.5 [13.5-28.0]), p<0.0001).

### External validation of key clinical associations for RALE scores

In an independent cohort of 415 COVID-19 inpatients (Table S4), we found that baseline RALE scores were significantly different between IMV (n=68) and SB (n=347, p<0.0001, Figure S6), we replicated the correlations between BMI and hypoxemia inferred by SpO2/FiO_2_ ratios, and validated the association between baseline RALE scores with 90-day mortality, with non-survivors having markedly higher RALE scores than survivors (p<0.0001, Figure S6).

## Discussion

Our study utilized the RALE scoring system to examine the radiographic heterogeneity of COVID-19 pneumonia among inpatients with a wide spectrum of clinical severity. With a systematic approach supported by a dedicated software, we demonstrated that RALE scoring is a learnable skill for clinicians, relatively easy to use, with excellent inter-rater agreement following appropriate training. We demonstrated that technical aspects of image quality and radiographic penetration impact RALE score assignments. Among inpatients with COVID-19, RALE scores were reflective of disease severity by level of respiratory support, significantly associated with patient-level pre-morbid covariates (such as age, BMI, history of COPD), correlated with respiratory dysfunction parameters (mechanics and gas exchange in IMV patients), were significantly associated with the adverse hyper-inflammatory subphenotype of host responses, and shown to be prognostic of survival and discharge destination among survivors.

To study the reproducibility of RALE scoring and obtain a reliable database of radiographic assessments by expert reviewers, our team created the *Pulmo-Annotator* software, which allowed for stable storage of images/scores on a cloud-based platform with parallel scoring from many individual reviewers. The *Pulmo-Annotator* capacities allowed us to study in depth technical aspects of image quality/penetration on resultant RALE scores, as well as reviewer-related sources of variation. We were able to easily identify sporadic discordant scores or systematic patterns of deviation by reviewer, provide iterative feedback and optimize inter-rater reliability. Our exercise showed that RALE scoring is a trainable skill but requires a systematic mechanism to accomplish high inter-rater agreement. With an expansive database of expert-annotated RALE scores and image attributes, RALE scoring may also become machine-learnable, which could transform the speed and scale of radiographic severity assessment in healthcare applications. There are multiple ongoing efforts in the field of machine learning for chest radiography^19^, but any type of sophisticated model will require high-quality image annotations by clinical experts - as pursued in our study - to generate valid predictions.

We found that pre-morbid demographic variables were significantly associated with RALE scores at time of hospitalization. Among IMV patients, those with possible indicators of frailty (such as older patients or nursing home residents) had significantly lower RALE scores, suggestive that such patients required a lower burden of acute respiratory illness to end up on IMV. Similarly, patients with COPD had lower RALE scores, perhaps also indicative of their limited physiologic reserve as well as the anatomical emphysema accounting for increased radiographic lucency. On the other hand, patients with higher BMI had higher RALE scores, which may reflect both the known association of obesity with COVID-19 severity,^20^ as well as diminished lung volumes and increased radiographic density from extra-thoracic soft tissue. Therefore, such pre-morbid variables need to be accounted in analyses of radiographic indices with clinical endpoints.

We studied a large sample of 425 inpatients with a wide spectrum of COVID-19 severity, as illustrated by the range of WHO scale from 4 to 9 at timing of CXR and demonstrated a stepwise increase of RALE scores by levels of respiratory support. We demonstrated significant associations of RALE scores not only with clinical severity, but also with detailed metrics of pulmonary physiology (mechanics and gas exchange) as well as administered therapies used for the most severely ill patients with COVID-19 pneumonia. We validated our observations in an independent cohort of COVID-19 inpatients enriched for non-intubated patients. Of note, we observed a temporal correlation of RALE scores in IMV patients with the time spent from onset of the pandemic, i.e. patients enrolled in 2021 having higher RALE scores than patients in the first waves of the pandemic in 2020. This temporal observation may reflect different population demographics (more frail patients hospitalized in 2020), evolving practices around initiation of IMV (more conservative criteria used as the pandemic progressed, and thus only sicker patients being intubated), or true, worse lung injury from emergent SARS-CoV-2 variants.

We detected novel associations of RALE scores with biomarkers of host innate immune response (IL-6 and sTNFR1) and lung epithelial injury (sRAGE) in IMV patients. The significant correlation between sRAGE levels and RALE scores validates previous findings^5,21–23^, but the newly detected associations with innate immunity biomarkers and the hyper-inflammatory subphenotype in both IMV and SB patients is suggesting that radiographic severity is not only representative of accumulated lung injury by the time of CXR, but also indicative of ongoing inflammatory damage. Our findings suggest that radiographic severity assessments in severe pneumonia and ARDS may offer further insights in ongoing efforts to better characterize and understand the biological and clinical heterogeneity of such complex syndromes, and RALE scoring is an accessible tool for such purposes.

With a larger sample size than previous studies^7,8,10,9,24–28^, and a systematic method supported by dedicated software, we validated the prognostic value of baseline RALE scores on clinical outcomes. Notably, RALE scores were predictive of 60-day survival even after adjustment of possible confounders (age, sex, history of COPD and BMI), which we chose to adjust for given their significant associations with RALE scores and known impact on COVID-19 outcomes. Nonetheless, when examined within each subgroup of levels of respiratory support (SB, IMV and ECMO), we did not find a significant prognostic effect of baseline RALE scores. Recent studies have shown that rising RALE scores on follow-up CXRs carry prognostic value in COVID-19^17^, and we had previously shown that declining RALE scores in patients with non-COVID ARDS were associated with liberation from mechanical ventilation^6^. Thus, although baseline RALE scores capture important cross-sectional parameters of clinical severity, reliable prognostication or assessment of treatment response may require longitudinal scoring of radiographic severity in the early period of hospitalization.

Our study has several limitations. For logistical/feasibility reasons, we analyzed only baseline CXRs from a total of 840 COVID-19 inpatients, and thus could not determine the trajectories of radiographic evolution that may offer important prognostic information. We analyzed biospecimens only from two inpatient cohorts (ALIR and COVID-INC) and therefore our biomarker analyses may have had limited statistical power to detect additional significant associations. We utilized portable CXR images obtained as part of routine medical care, and did not standardize image acquisition protocols for this study. Nonetheless, the analyzed dataset of images is representative of clinical practices in two major hospital systems and results are likely further generalizable.

CXRs represent the most used radiographic modality for diagnosis, monitoring severity and response to treatment among hospitalized patients with pneumonia. Although inferior in resolution and dimensionality compared to CT imaging, CXRs expose patients to substantially lower radiation dose, they are more rapid, cheaper, easily accessible and repeatable, and can be used in low resource care settings. Current clinical practice involves qualitative or implicit interpretations of CXRs, e.g. by narrative descriptions of densities (*focal, patchy* or *diffuse*) or qualifiers of progression (*improved* or *worse*). Such subjective, non-specific assessments are not reliable for objective evaluation of radiographic severity. Consequently, standard clinical practices fail to capitalize on objective imaging data provided by the most widely used modality. Our reproducible method for RALE scoring assessments offers a tool for thorough, quantitative study of radiographic severity.

With the wide availability of CXR imaging among hospitalized patients with COVID-19, incorporation of radiographic severity assessments into risk stratification may provide improved patient-level guidance on prognosis and treatment allocation.

## Supporting information

Supplemental Sheet

## Data Availability

All data produced in the present work are contained in the manuscript

## Acknowledgment

The authors would like to thank Olivia Glotfelty-Scheuering, a research librarian at UPMC Mercy Hospital, Manager of Library Services (MLIS), for her assistance in carrying out the literature search.

