## Supplemental Sheet for "Radiographic Assessment of Lung Edema (RALE) Scores are Highly Reproducible and Prognostic of Clinical Outcomes for Inpatients with COVID-19"

#### Expanded Methods

Discovery dataset: We analyzed data obtained from hospitalized patients with COVID-19, who were enrolled from April 2020 through October 2021 in one of three independent cohort studies within the UPMC Health System:

a. **The Acute Lung Injury Registry (ALIR) and Biospecimen Repository**. ALIR is a prospective cohort study of critically ill patients hospitalized in ICUs at UPMC Presbyterian/Shadyside and UPMC East hospitals. In this study, we included adult patients (18-90 years of age) diagnosed with COVID-19 based on respiratory symptoms, hypoxemia, and confirmatory testing via nasopharyngeal or lower respiratory tract (LRT) quantitative polymerase chain reaction (qPCR) for SARS-CoV-2 RNA. We enrolled subjects following admission to the ICU and obtained informed consent from the patients or their legally authorized representatives under the study protocol STUDY19050099 approved by the University of Pittsburgh Institutional Review Board (IRB). Upon enrollment, we collected blood biospecimens for centrifugation and separation of plasma and other blood constituents, which were stored in -80C until experiments.
b. **The COVID INpatient Cohort (COVID-INC).** COVID-INC is a prospective cohort study of moderately ill inpatients with COVID-19 hospitalized in dedicated inpatient wards at UPMC Presbyterian/Shadyside hospitals, as well as critically ill patients admitted to the ICU at UPMC East hospital. We enrolled adult patients (18-90 years of age) diagnosed with COVID-19 based on respiratory symptoms, hypoxemia and confirmatory testing via nasopharyngeal or LRT SARS-CoV-2 qPCR. We obtained consent from the patients or their legally authorized representatives under the study protocol STUDY20040036 approved by the University of Pittsburgh IRB. Upon enrollment, we collected blood biospecimens for centrifugation and separation of plasma and other blood constituents, which were stored in -80C until experiments.
c. **The Prognostication for COVID-19 Patients Admitted to Intensive Care Units at UPMC Pinnacle (PROCOPI) study.** The PROCOPI study is a retrospective cohort study of critically ill patients with COVID-19 hospitalized in ICUs at UPMC Pinnacle hospitals. Under a minimal risk study protocol (20E059) approved by the UPMC Pinnacle IRB, we performed a retrospective chart review and collected data from the electrical medical record (EMR) of patients with COVID-19 (diagnosed by nasopharyngeal or LRT SARS-CoV-2 qPCR) on IMV.

Validation cohort: We obtained admission CXRs from 415 COVID-19 inpatients hospitalized within 18 different clinical sites of the Cleveland Clinic systems (CCS) from March 2020 – October 2020. We collected clinical data from electronic medical records on demographics, comorbidities, physiologic and laboratory variables under an exempt review protocol (FLA 20-038) as previously described[1]. We classified patients into SB and IMV groups based on the type of respiratory support by the timing of the CXR. RALE scoring was performed by an independent set of investigators within the CCS.

Clinical data collection: For all subjects enrolled in any of the three derivation cohorts, we retrieved a single portable CXR image at a baseline timepoint, defined as following: i) day of hospital admission for the non-ICU patients of the COVID-INC cohort, ii) day of ICU admission for non-intubated, spontaneously breathing critically ill patients (ALIR and COVID-INC cohorts), iii) day of intubation for mechanically ventilated patients (ALIR, COVID-INC and PROCOPI cohorts). We stored CXR images in *.jpeg* format on a password protected HIPAA compliant institutional cloud and uploaded them in batches in the *Pulmo-Annotator* software. From the EMR, we extracted data on demographics, comorbid conditions, vital signs and laboratory test results at the time of baseline CXR. Based on the level of respiratory and other organ failure support, we scored each patient's severity of illness at baseline according to the 10-point ordinal scale of the World Health Organization (WHO). We broadly classified respiratory support in three categories at baseline: i) **SB** patients, i.e., not intubated subjects on various levels of oxygenation support, ii) **IMV**, intubated subjects in the ICU, and iii) **ECMO**, i.e., intubated subjects in the ICU on ECMO support at time of CXR. From IMV patients, we also collected detailed physiologic data from physician-set ventilatory parameters and obtained measurements, including Tidal Volume (expressed in ml per kg of ideal body weight [IBW]), Positive End Expiratory Pressure (PEEP), Fraction of Inspired Oxygen ($FiO_2$), Plateau Pressure, Driving Pressure, PaO2/FiO2 ratio and Ventilatory Ratio, as previously described[2,3]. For all patients, we recorded administered therapies for COVID-19 (steroids, tocilizumab and remdesivir), as well as adjunctive therapeutics for hypoxemic respiratory failure (prone positioning, neuromuscular blockade, and ECMO). We recorded clinical endpoints across a timeline of COVID-19 course starting from date of symptom onset to dates of hospital/ICU admission, dates of intubation/extubation for IMV subjects, date of discharge and destination for survivors, and date of death for non-survivors by 60 days from date of CXR. Time to event for clinical outcomes were calculated from the date of CXR till the date of the relevant event.

RALE scoring: We performed RALE score assessments by ≥2 independent reviewers per image with the *Pulmo-Annotator* software (Figure 1). Following a series of iterative training sessions on RALE scoring (~ 20 CXRs per reviewer), each trained reviewer was provided access to the *Pulmo-Annotator* software, with CXR sets divided in random batches of CXRs (range 25-90) and reviewers blinded to clinical data for each CXR. For each image, each reviewer first determined the horizontal level of the first branch of the left main bronchus and a vertical mid-point of the vertebral column, which the software used to divide the CXR into four quadrants (Right upper quadrant [RUQ], left upper quadrant [LUQ], right lower quadrant [RLQ] and left lower quadrant [LLQ]). Each reviewer then assessed the radiographic penetration (by determining the lowest level of visible vertebral bodies across mediastinal structures), overall image quality (subjective assessment as good or poor), presence of endotracheal tube, presence of right or left lung atelectasis (yes, no or possible), and then scored the most dense radiographic opacity in each quadrant by extent (scores of 0 for none, 1 for < 25%, 2 for 25–50%, 3 for 50–75% and 4 for > 75% of quadrant involved) and density (scores of 1 for hazy, 2 for moderate and 3 for dense consolidation). Each quadrant's score was automatically calculated as the product of extent*density, and then all four quadrant scores were summed for a final RALE score (ranging from 0-48)[4]. Following a first iteration of RALE scoring for each CXR batch by at least two reviewers, we reviewed all available data, examined distributions of scores and variables by each reviewer, and determined CXRs with large discrepancies between reviewers (a difference of score of ≥ 2 in any quadrant extent or density, or ≥15 RALE score difference). The reviewers then held joint sessions to review these CXRs and understand sources of disagreement, and then performed independent repeat RALE score assessments. Each CXR was reviewed by at least one senior reviewer (GDK, board certified physician in Pulmonary and Critical Care Medicine with extensive experience in RALE scoring and NAY, senior Internal Medicine Resident who was extensively trained by GDK on RALE scoring). We used the RALE scores and annotated variables from the second iteration in quantitative analyses.

Plasma biomarkers: We measured plasma biomarkers of injury and inflammation with custom-made Luminex panels as previously described[5], for available baseline samples from the ALIR and COVID-INC cohorts. We measured eight biomarkers of innate immune responses (interleukin [IL]-6, IL-8, soluble tumor necrosis factor receptor 1 [sTNFR1], suppression of tumorigenicity-2 [ST-2]), epithelial injury (soluble receptor of advanced glycation end-products [sRAGE]), endothelial injury (angiopoietin-2 [Ang2]) and host-response to bacterial infections (procalcitonin and pentraxin-3). From available biomarker values, we classified subjects into a hyperinflammatory vs. hypoinflammatory subphenotype by using predicted probabilities for subphenotype classifications from a published parsimonious logistic regression model utilizing IL-6, sTNFR1 and bicarbonate[6]. In a random subset of plasma samples, we quantified circulating levels of SARS-CoV-2 RNA by quantitative PCR, as previously described[7,8].

Statistical Analyses: We performed non-parametric comparisons for continuous (described as median and interquartile range – IQR) and categorical variables between clinical groups (Wilcoxon and Fisher's exact tests, respectively). We examined for inter-reviewer agreement on RALE scores with Bland-Altman plots pre- and post-feedback sessions, and quantitatively by measuring inter-reviewer correlations and Intraclass Correlation Coefficients (ICC) in two-way random-effects models (rated by ICC as poor [<0.50], moderate [0.50–0.75], good [0.75–0.90], and excellent [>0.9]). For categorical variables on CXR assessments, we quantified inter-reviewer agreement with Cohen's kappa statistics (rated as fair [0.21–0.40], moderate [0.41–0.60], substantial [0.61–0.80] and perfect [0.81–1.00]). We examined correlations of continuous variables with Pearson correlation test. We fit proportional hazards models to examine the statistical significance of baseline RALE scores on 60-day survival or time-to-liberation from IMV. We performed all analyses with the R software and a p-value of <0.05 was deemed statistically significant.

### Tables

Table S1: Baseline characteristics of enrolled subjects per UPMC cohort.

| | ALIR (n = 154) | COVID-INC (n = 138) | PROCOPI (n = 133) | p-value |
|---|---|---|---|---|
| **Demographics** | | | | |
| Age, Years | 59.5 [49.4, 70.6] | 63.0 [54.0, 70.8] | 68.0 [60.0, 76.0] | <0.01 |
| Body Mass Index | 33.5 [27.5, 38.6] | 30.3 [26.2, 37.1] | 32.4 [28.5, 36.9] | 0.11 |
| Gender, Male | 99 (64.3%) | 58 (42.0%) | 97 (72.9%) | <0.01 |
| Race, Whites | 122 (79.2%) | 109 (79.0%) | 92 (69.2%) | <0.01 |
| Never Smoker | 47 (44.3%) | 69 (52.3%) | 58 (43.6%) | 0.3* |
| Resident of Nursing Facility | 16 (15.1%) | 14 (10.1%) | 20 (15.2%) | 0.39* |
| Diabetes Mellitus | 61 (39.6%) | 54 (39.1%) | 66 (49.6%) | 0.14 |
| Chronic Obstructive Lung Disease | 23 (14.9%) | 24 (17.4%) | 27 (20.3%) | 0.49 |
| Congestive Cardiac Failure | 21 (13.6%) | 18 (13.0%) | 20 (15.0%) | 0.89 |
| **Plasma biomarkers (pg/ml)** | | | | |
| IL-6 | 76.5 [17.3, 263.8] | 11.4 [4.6, 26.2] | NA [NA, NA] | <0.01 |
| IL-8 | 22.8 [15.0, 40.1] | 12.6 [6.2, 21.2] | NA [NA, NA] | <0.01 |
| Ang2 | 3949.3 [2412.8, 8179.8] | 2838.7 [1752.0, 4228.7] | NA [NA, NA] | <0.01 |
| Procalcitonin | 414.2 [145.9, 1508.5] | 96.6 [63.4, 249.8] | NA [NA, NA] | <0.01 |
| ST2 | 148584.4 [72120.4, 254988.8] | 87770.0 [45738.7, 190531.0] | NA [NA, NA] | <0.01 |
| Pentraxin-3 | 8211.2 [4126.7, 19479.2] | 5318.4 [2350.6, 10441.2] | NA [NA, NA] | <0.01 |
| sRAGE | 5576.6 [2581.2, 11477.9] | 3589.6 [2131.9, 8455.6] | NA [NA, NA] | 0.05 |
| sTFNR1 | 5098.6 [3397.6, 10142.5] | 3166.0 [2279.1, 5021.8] | NA [NA, NA] | <0.01 |
| Hyperinflammatory Subphenotype | 19 (16.1%) | 0 (0%) | NA (NA) | <0.01* |
| **Radiographic parameters** | | | | |

| | | | | |
|---|---|---|---|---|
| RALE Score | 28.0 [20.9, 35.9] | 19.0 [13.3, 24.7] | 26.0 [19.5, 32.8] | <0.01 |
| Lower Quadrants RALE Score | 17.5 [14.0, 22.0] | 13.2 [9.0, 16.0] | 16.0 [12.0, 20.0] | <0.01 |
| Upper Quadrants RALE Score | 11.0 [6.2, 14.0] | 6.0 [3.5, 9.0] | 9.0 [7.0, 13.2] | <0.01 |
| Time of CXR from symptom onset, days | 8.0 [4.0, 13.0] | 7.0 [3.0, 10.0] | 10.0 [6.0, 16.0] | <0.01 |
| **Clinical outcomes** | | | | |
| In Hospital Mortality | 64 (41.6%) | 14 (9.5%) | 125 (94.0%) | <0.01 |
| Discharged To | | | | |
| Home | 36 (23.4%) | 95 (68.8%) | 0 (0.0%) | <0.01 |
| In Patient Rehab | 19 (12.3%) | 0 (0.0%) | 0 (0.0%) | <0.01 |
| LTAC | 14 (9.1%) | 5 (3.6%) | 5 (3.8%) | <0.01 |
| SNF | 21 (13.6%) | 25 (18.1%) | 3 (2.3%) | 0.444 |

Continuous variables are reported in median [interquartile range]. Nominal variables are reported in n (%).

* = we only included patients with available clinical data or research biomarkers in analysis. Patients with unavailable data were excluded.

Definition of abbreviations: CXR = Chest X-Ray, PEEP = Positive End Expiratory Pressure, PaO2 = Partial Pressure of Oxygen, FiO2 = Fraction of Inspired Oxygen, IL = Interleukin, Ang2 = Angiopoietin-2, ST2 = Suppression of Tumorigenicity-2, sRAGE = Soluble Receptor of Advanced Glycation End-Products, NMBA = Neuromuscular Blocking Agents, ECMO = Extracorporeal Membrane Oxygenation, IPR = Inpatient Rehab, LTAC = Long Term Acute Care, and SNF = Skilled Nursing Facility.

Table S2: Pre-post feedback inter-rater agreement for RALE score

| | Pre-feedback | | | Post-feedback | | |
|---|---|---|---|---|---|---|
| **Validator** | **Number of CXR** | **ICC2k (lower – upper bound)** | **p-value** | **Number of CXR** | **ICC2k (lower – upper bound)** | **p-value** |
| **2.0.1** | 95 | 0.90 (0.83-0.94) | <0.001 | 95 | 0.96 (0.94-0.98) | <0.001 |
| **2.0.2** | 63 | 0.83 (0.19-0.94) | <0.001 | 63 | 0.88 (0.47-0.96) | <0.001 |
| **2.1.0** | 25 | 0.92 (0.82-0.96) | <0.001 | 25 | 0.96 (0.92-0.98) | <0.001 |
| **2.1.1** | 25 | 0.87 (0.71-0.94) | <0.001 | 25 | 0.97 (0.92-0.99) | <0.001 |
| **2.1.2** | 47 | 0.67 (-0.21-0.89) | <0.001 | 47 | 0.86 (-0.08-0.96) | <0.001 |
| **2.1.3** | 47 | 0.85 (0.64-0.93) | <0.001 | 47 | 0.94 (0.85-0.97) | <0.001 |
| **2.1.4** | 91 | 0.85 (0.77-0.90) | <0.001 | 98 | 0.94 (0.91-0.96) | <0.001 |
| **2.1.5** | 41 | 0.80 (0.35-0.92) | <0.001 | 41 | 0.85 (0.32-0.94) | <0.001 |
| **Combined** | 434 | 0.86 (0.83-0.88) | <0.001 | 441 | 0.93 (0.92-0.95) | <0.001 |

Table S3: Cohen's Kappa for 2 Raters for radiographic variables

| Variable | Kappa | p-value |
|---|---|---|
| **Intubation Status** | 1.00 | <0.001 |
| **Quality of Image** | 0.60 | <0.001 |
| **Left Atelectasis** | 0.21 | <0.001 |
| **Right Atelectasis** | 0.21 | <0.001 |
| **X-ray Penetration** | 0.18 | <0.001 |

#### Figures

**Figure S1: CXR image variables and RALE scores:**

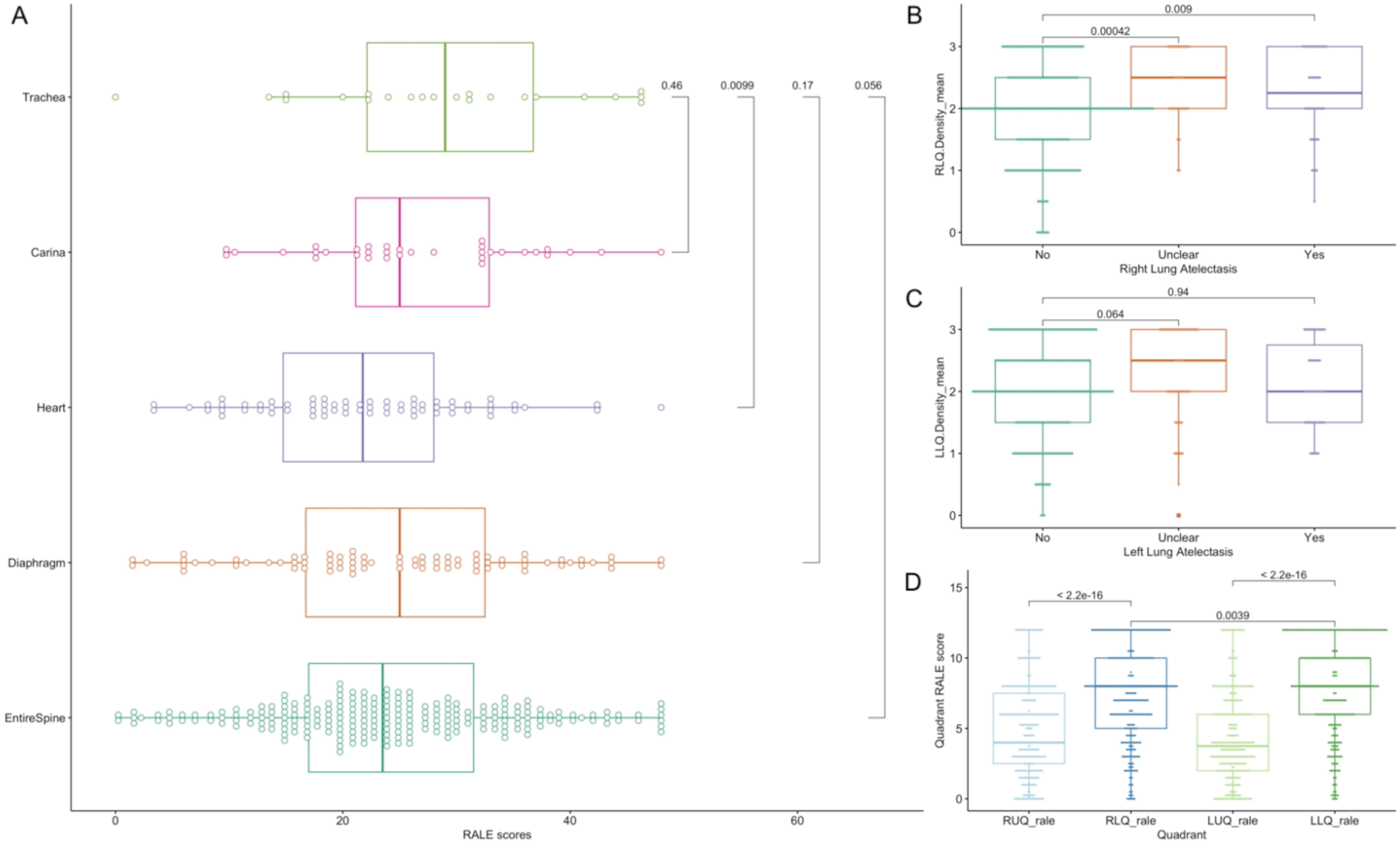

A: Radiographic penetration and RALE scores. Y-axis denotes the lowest anatomical level of mediastinal structures behind which the vertebral bodies were visible. Under-penetrated CXRs (vertebral bodies were visible only behind the trachea) had higher median RALE scores compared to CXRs with visible vertebral bodies behind the heart ($p<0.01$). No significant difference was noted among other levels of CXR penetration. B: The absence of right lung atelectasis was associated with lower mean RLQ density score compared to CXRs with present or possibly present atelectasis ($p<0.01$). C: Left lung atelectasis was not significantly associated with differences in LLQ density scores. D: The lower quadrants (right and left) had much higher quadrant scores compared to their corresponding upper quadrants (right and left, respectively, $p<0.0001$), whereas LLQ scores were statistically significantly higher than RLQ ones ($p<0.01$, Fig S1D).

Definition of abbreviations: CXR = Chest X-Ray, RUQ = Right Upper Quadrant, RLQ = Right Lower Quadrant, LUQ = Left Upper Quadrant, and LLQ = Left Lower Quadrant

**Figure S2: RALE scores by enrollment cohort and by timing of enrollment during the pandemic, stratified by level of respiratory support.**

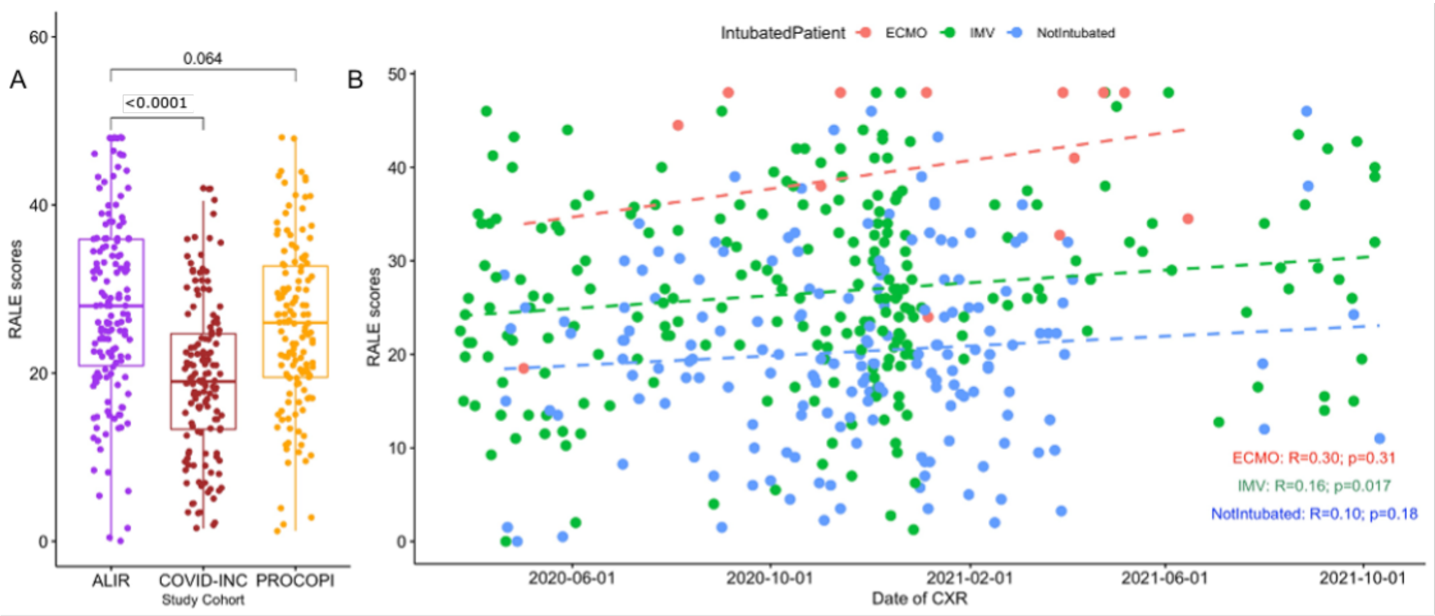


A: Median [Interquartile range] of RALE score was lower in the COVID-INC (19.0 [13.3, 24.7]) compared to the ALIR (28.0 [20.9, 35.9]) and PROCOPI (26.0 [19.5, 32.8], p<0.0001) cohorts. There was no significant difference between ALIR and PROCOPI cohorts (p = 0.064). B: Across the timeframe for enrolment, RALE scores were weakly correlated with the time spent from the onset of the pandemic in IMV patients (R = 0.16, p = 0.017). No significant correlation was seen for ECMO or SB patients.

Definition of abbreviations: ALIR = The Acute Lung Injury Registry and Biospecimen Repository, COVID-INC = The COVID Inpatient Cohort, PROCOPI = The Prognostication for COVID-19 Patients Admitted to Intensive Care Units at UPMC Pinnacle study, ECMO = Extracorporeal Membrane Oxygenation, and IMV = Invasive Mechanical Ventilation

**Figure S3: RALE score associations with demographic categorical variables.**

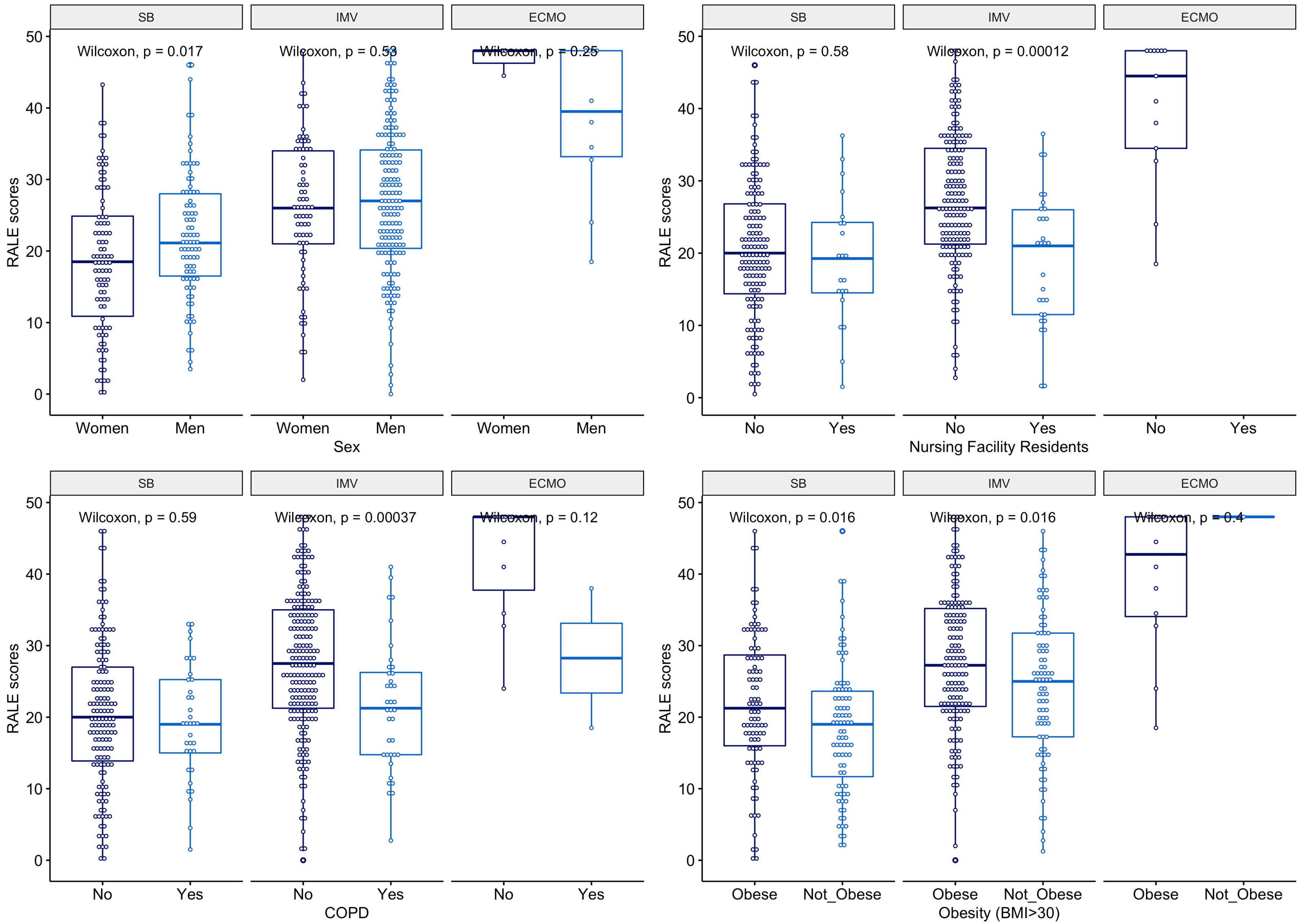

This figure shows the median RALE scores for basic demographics among the three groups of respiratory support. For SB patients, male and obese patients had higher median RALE score (p = 0.017 and p = 0.016, respectively). For IMV patients, obese patients had higher median RALE score (p = 0.016), whereas SNF residents and patients with COPD had lower median RALE scores (p<0.001).

Definition of abbreviations: SB = Spontaneously Breathing, IMV = Invasive Mechanical Ventilation, ECMO = Extracorporeal Membrane Oxygenation, SNF = Skilled Nursing Facility, and COPD = Chronic Obstructive Lung Disease.

**Figure S4: RALE score correlations with baseline continuous variables.**

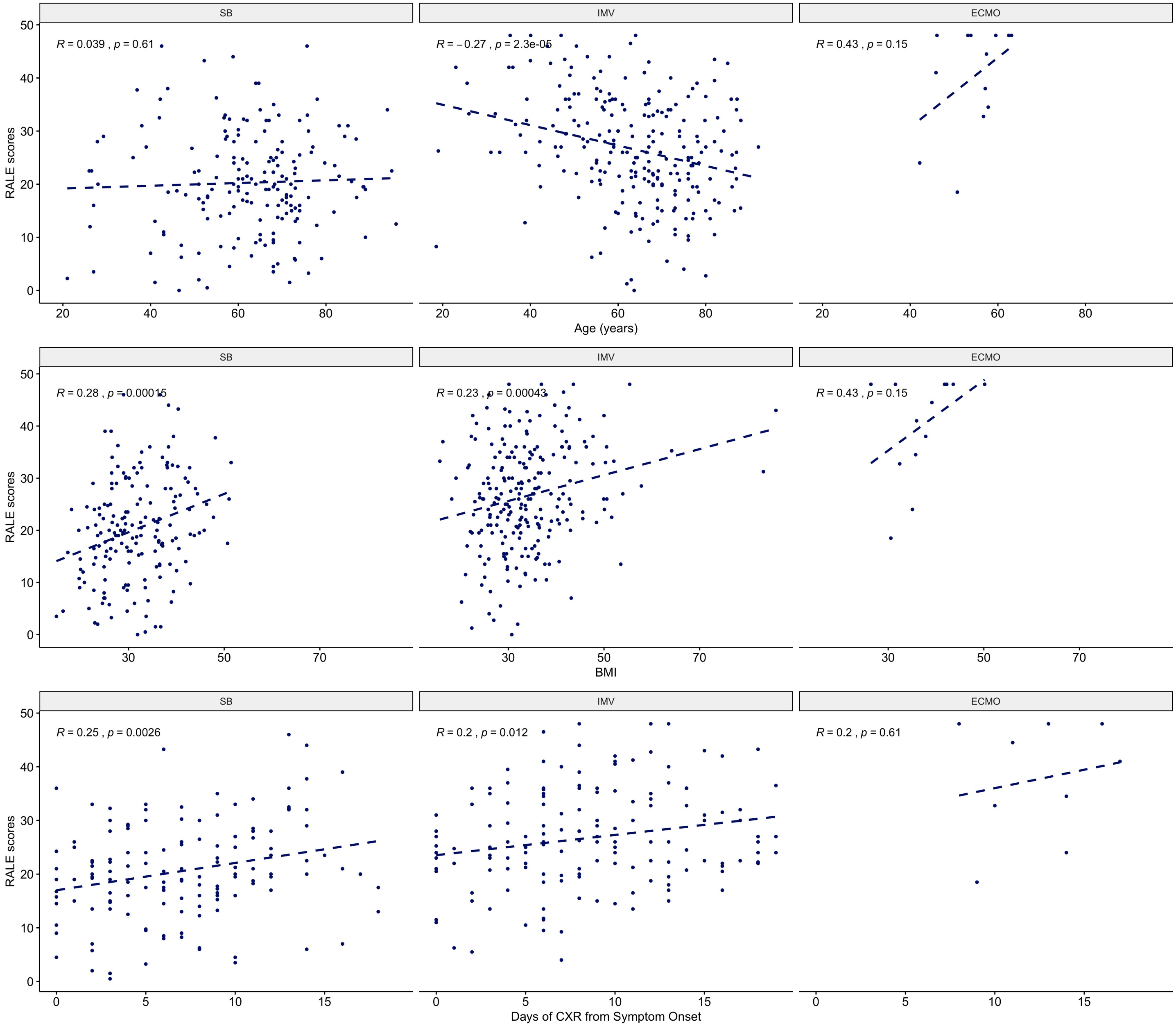

For non-intubated patients, RALE score was correlated with BMI (r = 0.28, p <0.001) and time from symptom onset (r = 0.25, p = 0.0026). For IMV patients, RALE score was correlated with BMI (r = 0.23, p <0.001) and time from symptom onset (r = 0.2, p = 0.012) and inversely correlated with age (r = -0.27, p<0.0001).

Definition of abbreviations: SB = Spontaneously Breathing, IMV = Invasive Mechanical Ventilation, ECMO = Extracorporeal Membrane Oxygenation, BMI = Body Mass Index, and CXR = Chest X-Ray

**Figure S5: RALE score association with adjunctive therapeutics for hypoxemic respiratory failure and COVID-19 specific treatments.**

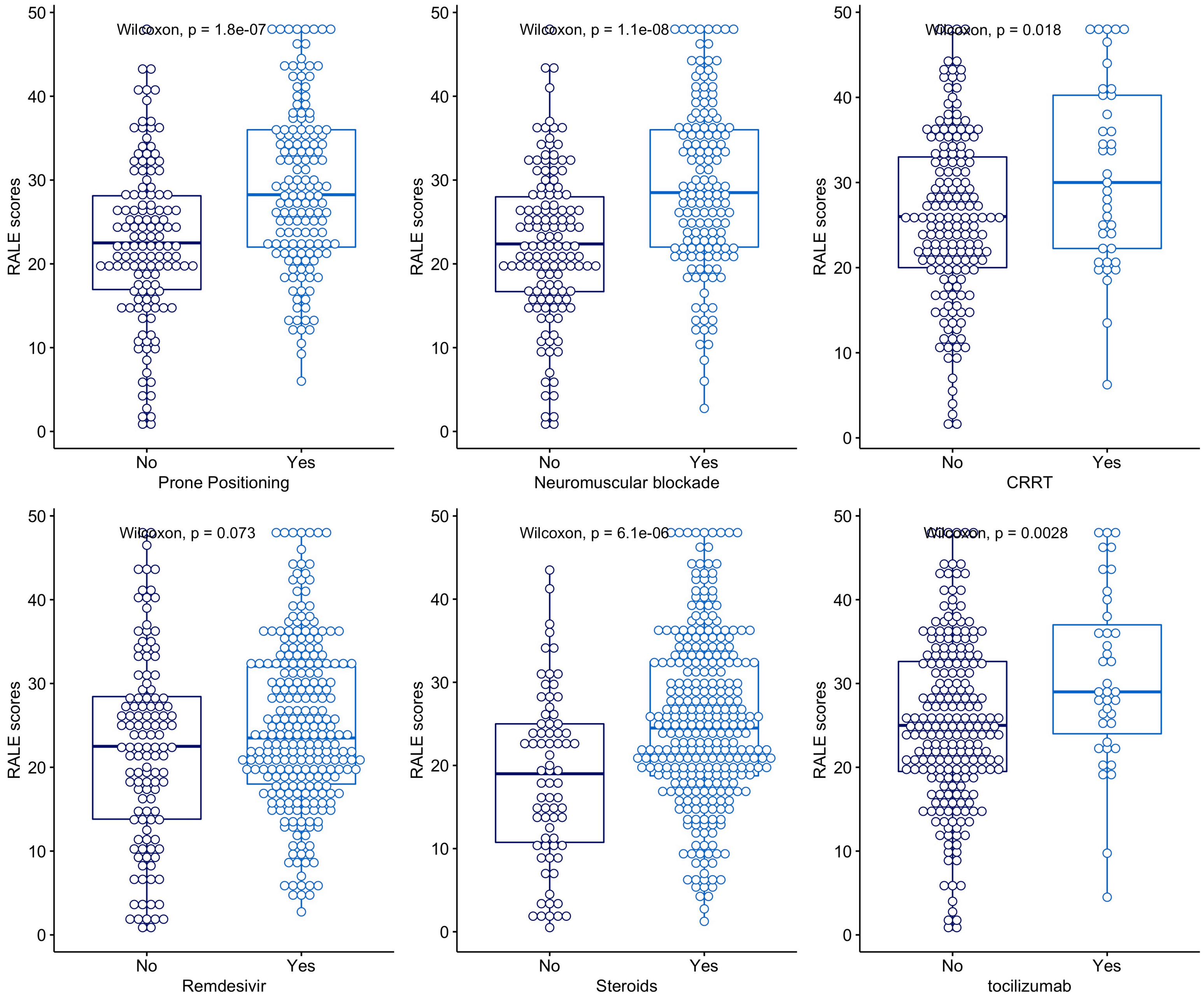

Patients who received neuromuscular blockade therapy, prone positioning, continuous renal replacement therapy, steroids or tocilizumab had higher RALE scores compared to their untreated counterparts.

**Figure S6: Validation results from the Cleveland Clinic Foundation Cohort**

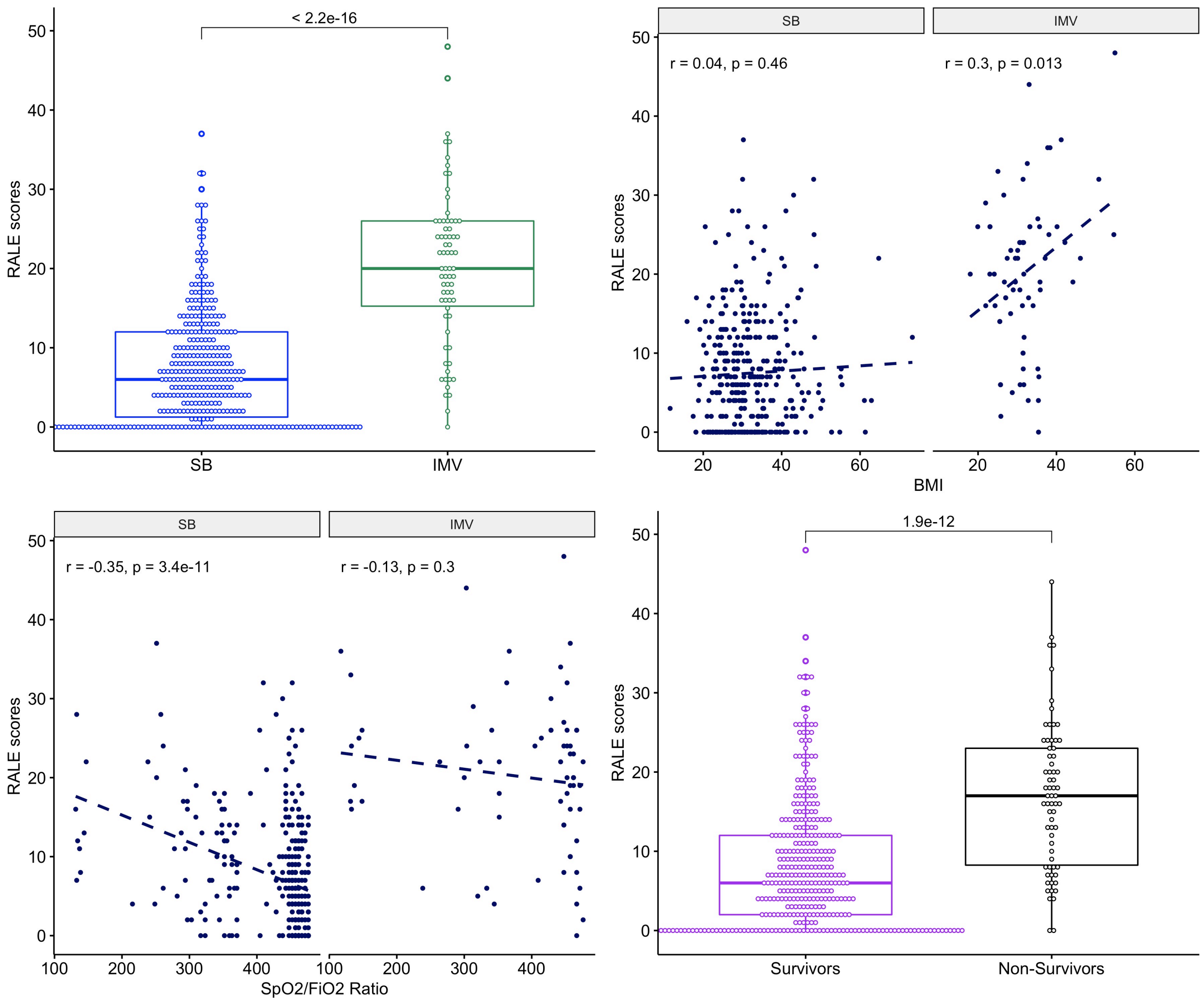


Among inpatients from the Cleveland Clinic Foundation hospital system, IMV patients had higher RALE score compared to SB patients. For IMV patients, RALE scores were significantly correlated with BMI. For SB patients, RALE score inversely corelated with worse $SpO_2/FiO_2$ ratio. RALE scores were also higher among non-survivors.

Table S4: Cleveland Clinic Foundation cohort baseline characteristics

| | IMV (n = 68) | SB (n = 347) | P-value |
|---|---|---|---|
| **Demographics** | | | |
| **Age, Years** | 68.3 [60.7, 75.4] | 60.7 [48.6, 72.9] | <0.01 |
| **Body Mass Index** | 31.4 [27.1, 35.4] | 30.6 [26.3, 36.2] | 1.00 |
| **Sex, Male** | 41 (60.3%) | 167 (48.1%) | 0.09 |
| **Race, White** | 31 (45.6%) | 132 (38.0%) | 0.56 |
| **Never Smoker** | 37 (54.4%) | 204 (58.8%) | 0.65 |
| **Diabetes Mellitus** | 30 (44.1%) | 101 (29.2%) | 0.02 |
| Chronic Obstructive Lung Disease | 24 (35.3%) | 94 (27.2%) | 0.23 |
| **Radiographic parameters** | | | |
| **RALE Score** | 20.0 [16.0, 26.0] | 6.0 [2.0, 12.0] | <0.01 |
| **Clinical outcomes** | | | |
| **Mortality at 90 Days** | 42 (61.8%) | 26 (7.5%) | <0.01 |

Continuous variables are reported in median [interquartile range]. Nominal variables are reported in n (%).

Definition of abbreviations: IMV = Invasive Mechanical Ventilation, SB = Spontaneously Breathing, COPD = Chronic Obstructive Pulmonary Disease

STROBE Statement—checklist of items that should be included in reports of observational studies

| | Item No. | Recommendation | Page No. | Relevant text from manuscript |
|---|---|---|---|---|
| **Title and abstract** | 1 | (*a*) Indicate the study's design with a commonly used term in the title or the abstract | 3 | |
| | | (*b*) Provide in the abstract an informative and balanced summary of what was done and what was found | 3 | We performed independent … Line 61-77 |
| **Introduction** | | | | |
| Background/rationale | 2 | Explain the scientific background and rationale for the investigation being reported | 4-5 | Multiple risk stratification … Line 86-105 |
| Objectives | 3 | State specific objectives, including any prespecified hypotheses | 4-5 | In this study, we investigated … Line 106-111 |
| **Methods** | | | | |
| Study design | 4 | Present key elements of study design early in the paper | 5 | We analyzed data obtained … Line 114-116 |
| Setting | 5 | Describe the setting, locations, and relevant dates, including periods of recruitment, exposure, follow-up, and data collection | 5 | We analyzed data obtained … Line 114-155 |
| Participants | 6 | (*a*) *Cohort study*—Give the eligibility criteria, and the sources and methods of selection of participants. Describe methods of follow-up<br>*Case-control study*—Give the eligibility criteria, and the sources and methods of case ascertainment and control selection. Give the rationale for the choice of cases and controls<br>*Cross-sectional study*—Give the eligibility criteria, and the sources and methods of selection of participants | 5 | We analyzed data obtained … Line 114-127 |
| | | (*b*) *Cohort study*—For matched studies, give matching criteria and number of exposed and unexposed<br>*Case-control study*—For matched studies, give matching criteria and the number of controls per case | 5-6 | Clinical data collection … Line 128-136 |
| Variables | 7 | Clearly define all outcomes, exposures, predictors, potential confounders, and effect modifiers. Give diagnostic criteria, if applicable | 5-6 | Clinical data collection … Line 128-155 |
| Data sources/ measurement | 8* | For each variable of interest, give sources of data and details of methods of assessment (measurement). Describe comparability of assessment methods if there is more than one group | 5-6 | Clinical data collection … Line 128-155 |
| Bias | 9 | Describe any efforts to address potential sources of bias | 7 | Validation cohort … Line 165-169 |
| Study size | 10 | Explain how the study size was arrived at | 7 | We analyzed baseline CXRs … Line 174-176 |
| Quantitative variables | 11 | Explain how quantitative variables were handled in the analyses. If applicable, describe which groupings were chosen and why | 6-7 | Statistical Analyses … Line 156-164 |
| Statistical methods | 12 | (*a*) Describe all statistical methods, including those used to control for confounding | 6-7 | Statistical Analyses … Line 156-164 |

| | | | | |
|---|---|---|---|---|
| | | (*b*) Describe any methods used to examine subgroups and interactions | 6-7 | Statistical Analyses … Line 156-164 |
| | | (*c*) Explain how missing data were addressed | | No missing data |
| | | (*d*) *Cohort study*—If applicable, explain how loss to follow-up was addressed<br>*Case-control study*—If applicable, explain how matching of cases and controls was addressed<br>*Cross-sectional study*—If applicable, describe analytical methods taking account of sampling strategy | | No loss to follow up |
| | | (*e*) Describe any sensitivity analyses | 6-7 | Statistical Analyses … Line 156-164 |
| **Results** | | | | |
| Participants | 13* | (a) Report numbers of individuals at each stage of study—eg numbers potentially eligible, examined for eligibility, confirmed eligible, included in the study, completing follow-up, and analysed | 7 | Characteristics of enrolled … Line 173-176 |
| | | (b) Give reasons for non-participation at each stage | | 100% participation |
| | | (c) Consider use of a flow diagram | | Not done |
| Descriptive data | 14* | (a) Give characteristics of study participants (eg demographic, clinical, social) and information on exposures and potential confounders | 14-15 | Table 1 |
| | | (b) Indicate number of participants with missing data for each variable of interest | | No missing data |
| | | (c) *Cohort study*—Summarise follow-up time (eg, average and total amount) | | Not applicable to our study |
| Outcome data | 15* | *Cohort study*—Report numbers of outcome events or summary measures over time | *7-10* | *Results, tables and figures Line 172-253* |
| | | *Case-control study*—Report numbers in each exposure category, or summary measures of exposure | | |
| | | *Cross-sectional study*—Report numbers of outcome events or summary measures | | |
| Main results | 16 | (*a*) Give unadjusted estimates and, if applicable, confounder-adjusted estimates and their precision (eg, 95% confidence interval). Make clear which confounders were adjusted for and why they were included | 7-10 | *Results … Line 172-253* |
| | | (*b*) Report category boundaries when continuous variables were categorized | 7-10 | *Results … Line 172-253* |
| | | (*c*) If relevant, consider translating estimates of relative risk into absolute risk for a meaningful time period | | |
| Other analyses | 17 | Report other analyses done—eg analyses of subgroups and interactions, and sensitivity analyses | | |
| **Discussion** | | | | |
| Key results | 18 | Summarise key results with reference to study objectives | 10-11 | Discussion … Line 255-265 |
| Limitations | 19 | Discuss limitations of the study, taking into account sources of potential bias or imprecision. Discuss both direction and magnitude of any potential bias | 13 | Our study has several limitations … Line 320-326 |
| Interpretation | 20 | Give a cautious overall interpretation of results considering objectives, limitations, multiplicity of analyses, results from similar studies, and other relevant evidence | 10-13 | Discussion … Line 255-338 |
| Generalisability | 21 | Discuss the generalisability (external validity) of the study results | 11-13 | To study the reproducibility … |

| | | | | |
|---|---|---|---|---|
| | | | | Line 266-319 |
| **Other information** | | | | |
| Funding | 22 | Give the source of funding and the role of the funders for the present study and, if applicable, for the original study on which the present article is based | 2 | Funding information … Line 48-49 |

*Give information separately for cases and controls in case-control studies and, if applicable, for exposed and unexposed groups in cohort and cross-sectional studies.

**Note:** An Explanation and Elaboration article discusses each checklist item and gives methodological background and published examples of transparent reporting. The STROBE checklist is best used in conjunction with this article (freely available on the Web sites of PLoS Medicine at http://www.plosmedicine.org/, Annals of Internal Medicine at http://www.annals.org/, and Epidemiology at http://www.epidem.com/). Information on the STROBE Initiative is available at www.strobe-statement.org.
